# Labour Induction in low-risk women at 39 weeks of gestation: a Randomised trial in China (LIRIC) - Protocol of an open label, randomised controlled trial

**DOI:** 10.64898/2026.05.24.26354001

**Authors:** Huajing Gao, Junhua Shen, Danqing Chen, Ben W. Mol, Wensheng Hu, Zhaoxia Liang, Xiaoxia Bai, Xiujun Han, Jiajun Zhu, Hong Wang, Xiaozhen Liu, Chang Su, Yuan Chen, Ruopeng Weng, Yifeng Liu, Wentao Li, Dan Zhang

## Abstract

**Introduction:** The ARRIVE trial first demonstrated that elective induction of labour (IOL) at 39 weeks in low-risk pregnancies reduced the likelihood of caesarean section (CS) without compromising perinatal safety; however, the generalizability of these findings remains debated, leading to uncertainty in clinical practice. The LIRIC trial aims to evaluate whether 39-week elective IOL reduces CS rates compared with expectant management, while exploring its impact on infant neurodevelopment and multi-omics profiles.

**Methods and analysis:** This is a single-centre, open-label, randomized controlled trial in China. A total of 1,074 low-risk pregnant women (nulliparous or multiparous) will be randomly assigned (1:1 ratio) to either 39-week IOL or expectant management. The primary outcome is the caesarean section (CS) rate. Secondary outcomes include a composite of severe neonatal morbidity and perinatal mortality and infant neurodevelopmental scores (Bayley-4 and ASQ-3), among others. Data analysis will follow the Intention-to-Treat (ITT) principle. Biospecimen will be collected for metagenomic and metabolomic analyses, with results to be reported separately.

**Ethics and dissemination:** The protocol has been approved by the Ethics Committee of Women’s Hospital, School of Medicine, Zhejiang University. Informed consent will be obtained from all participants. Results will be disseminated via peer-reviewed journals, and standardized infant developmental reports will be provided to participants to enhance study benefit.

**Trial registration number NCT07082530**.

## Introduction

Induction of labour (IOL) is a medical intervention to start the labour process before its spontaneous onset. It is one of the most widely utilized obstetric interventions globally. A 2010 World Health Organization (WHO) survey estimated IOL rates with a global baseline of 9.6%(1), and rates have increased over the past decade, frequently exceeding 30% in high-income countries. Between 2016 and 2024, IOL rates rose from 24.9% to 34.5% in the U.S.(2), and from 28% to 30% in the U.K.(3). In Australia, the rate reached 32.7% in 2023, compared with 25.2% in 2010(4). In China, a large national sampling survey in 2016 reported IOL rates of 18.4% in nulliparous and 10.2% in multiparous women(5). These figures indicate a substantial and rising use of IOL across diverse obstetric settings.

Historically, IOL has been reserved for situations in which continuing pregnancy increased risk for the mother or the child, or in post-term pregnancies. However, the use of IOL has been gradually expanded to low-risk pregnancies to improve obstetric outcomes and perinatal outcomes(6). The optimal timing of IOL in low-risk pregnancies has been a subject of ongoing debate over the past decade. In 2019, the INDEX and SWEPIS randomized controlled trials (RCTs) demonstrated that IOL at 41 weeks significantly reduced perinatal mortality and adverse perinatal outcomes compared with expectant management until 42 weeks(7, 8). Following these findings, the WHO and various national guidelines now recommend IOL at 41 weeks for low-risk pregnancies(9-13).

Whether IOL should be offered before 41 weeks in low-risk pregnancies remain uncertain. The ARRIVE RCT found IOL at 39 weeks did not significantly alter risks of severe composite perinatal outcomes but did reduce caesarean rates(14). Subsequent meta-analyses suggested further benefits, including reduced risks of emergent CS rates and grade III/IV perineal lacerations(15). However, most existing evidence is derived from Western populations, leaving a critical gap in implementation for other populations whose clinical characteristics and obstetric outcomes may differ. Consequently, the WHO emphasizes the need for more robust evidence before endorsing widespread 39-week IOL(9), while current Chinese guidelines restrict 39-week IOL to cases with suspected macrosomia(10), despite increasing clinical demand. The WHO further acknowledges that decision-making regarding IOL between 39 and 41 weeks may be driven by maternal anxiety or personal preferences(9). These shifts necessitate further evaluation of the risks and benefits of 39-week IOL in low-risk pregnancies to strengthen the global evidence base.

Furthermore, the long-term impact of IOL at 39 weeks on offspring remains inconclusive, particularly with respect to neurodevelopment; previous studies have focused on school-age academic performance(16-18), which may not accurately reflect early neurodevelopment and is subject to multiple postnatal influences over time. Evidence on neurodevelopmental outcomes in early infancy is limited. Also, because prior trials did not collect sequential biological samples surrounding IOL, they were unable to investigate biomarkers or mechanistic pathways linked to clinical outcomes.

This study, LIRIC, aims to address these gaps by conducting an RCT in Chinese low-risk pregnancies. We will evaluate whether 39-week IOL reduces CS rates compared to expectant management and extend the follow-up to six months postpartum to assess early infant development. By integrating clinical outcomes with multi-omics analyses of biospecimens, LIRIC will explore potential biological pathways linking IOL to early-life outcomes and provide evidence to inform clinical decision-making.

## Methods

### Study Design and Setting

LIRIC is an open-label, single-centre, superiority RCT. The protocol report adheres to the Standard Protocol Items: Recommendations for Interventional Trials (SPIRIT) 2025 guidelines(19). The study is conducted at the Women’s Hospital, Zhejiang University School of Medicine, which serves as the trial sponsor and reported over 28000 deliveries in 2024. The study enrollment, treatment and assessment plan are summarised in Table 1. Ethical approval has been received from the Ethics Committee of Women’s Hospital, School of Medicine, Zhejiang University (approval date: 10 June 2025)

**Table 1.** Study schedule.

| Timepoint | Trial period |  |  |  |  |  |  |  |
| --- | --- | --- | --- | --- | --- | --- | --- | --- |
|  | Enrollment |  | Post-randomisation |  |  |  |  | Close-out |
|  | 37w-38w | 38w+4d to 39w+3d | Birth | 1w old | 3w old | 6w old | 3m old | 6m old |
| Enrollment |  |  |  |  |  |  |  |  |
| Eligibility screen | X | X |  |  |  |  |  |  |
| Informed consent |  | X |  |  |  |  |  |  |
| Randomisation |  | X |  |  |  |  |  |  |
| Baseline variables | X | X |  |  |  |  |  |  |
| Intervention or comparator |  |  |  |  |  |  |  |  |
| Intervention |  |  | X |  |  |  |  |  |
| Comparator |  |  | X |  |  |  |  |  |
| Assessments and follow-up |  |  |  |  |  |  |  |  |
| Primary outcome |  |  | X |  |  |  |  |  |
| Secondary outcomes |  |  | X |  |  | X |  | X |
| Biological samples |  |  | X | X | X | X | X | X |

### Participants

Potential participants will be screened for eligibility during routine 37-week antenatal visits. Obstetricians will identify women with low-risk pregnancy and provide study information. Interested individuals will be referred to a dedicated research coordinator for detailed counselling and eligibility screening.

#### Inclusion Criteria

1. Age ≥18 years.
2. Singleton pregnancy or twin pregnancy reduced to singleton before 14 weeks of gestation.
3. Gestational age between 38 weeks 4 days and 38 weeks 6 days at randomization.
4. Eligible for vaginal delivery and has a desire for vaginal birth.
5. Reliable gestational age determination (confirmed by methods below).
6. Maternal and fetal conditions assessed as low-risk by at least two senior obstetricians, with no indications requiring delivery before 41 weeks.
7. Ability to understand study information and provide informed consent.

#### Gestational Age (GA) Determination

- In vitro fertilisation and embryo transfer (IVF-ET): calculated based on the embryo transfer and embryo age at transfer.
- Spontaneous conceptions or those via ovulation induction or intrauterine insemination:
  1. If last menstrual period (LMP) is reliable and matches the first-trimester ultrasound estimate (±7 days), LMP is used; otherwise, GA is determined by the crown-rump length (CRL);
  2. If LMP is unreliable, GA is determined by the first ultrasound CRL measurement before 14 weeks 0 days.

#### Exclusion Criteria

1. First-trimester ultrasound estimate >13 weeks 6 days.
2. Planned induction before 41 weeks.
3. Planned cesarean delivery or contraindications to vaginal delivery.
4. Already delivered, in labor, or ruptured membranes at enrollment.
5. Placenta previa, vasa previa, placenta accreta, or placental abruption.
6. Contraindications to labour induction (e.g., cervical cancer, history of uterine rupture, genital tract malformations, abnormal fetal position, cord prolapse).
7. Active vaginal bleeding and the volume is greater than spotting.
8. History of cesarean delivery or uterine/cervical surgery.
9. Cervical cerclage in the current pregnancy.
10. Maternal comorbidities or pregnancy complications that deemed unsuitable for expectant management beyond 39 weeks (e.g., pregestational diabetes, gestational diabetes requiring insulin, hypertensive disorders, intrahepatic cholestasis of pregnancy)(20).
11. Fetal conditions not suitable for expectant management beyond 39 weeks (e.g., fetal death, major anomalies, growth restriction, macrosomia, anemia, oligohydramnios, polyhydramnios)(20).
12. Maternal infections or positive screenings for sexually transmitted pathogens or group B Streptococcus.
13. Planned delivery at a non-study facility.
14. Participation in another intervention study affecting delivery management.

### Randomization and blinding

Eligible participants will be randomly assigned to either the 39-week IOL group or the expectant management group in a 1:1 ratio using stratified block randomization with variable block sizes (2 or 4). Randomization will be stratified by maternal age (<35 or ≥35 years) and parity (nulliparous or parous). The randomization sequence will be generated in advance by an independent third-party provider and integrated into the Electronic Data Capture (EDC) system to ensure strict allocation concealment. The sequence will remain inaccessible to all researchers and participants. Following reconfirmation of eligibility and informed consent, certified researchers will perform the randomisation via the EDC platform between 38 weeks 4 days and 38 weeks 6 days of gestation. Once allocated, the record will be automatically locked.

Given the nature of the intervention, masking to participants and clinicians is not possible; however, to mitigate potential bias, the statistical analysts and pediatricians evaluating offspring outcomes at the follow-ups will remain masked to the group allocation.

### Intervention and comparator

#### Standard of Care

Clinical management is guided by standardized protocols for IOL and expectant management, which are disseminated to all participating obstetricians as clinical algorithms (Supplementary Fig. 1-3) during Study Initiation Visit (SIV). Subsequent medical decisions during labour, including the decision to proceed to CS, remain at the discretion of the attending physician based on real-time clinical requirements. Indications for CS include, but are not limited to, failed indication, fetal distress, labour arrest, or other obstetric emergencies.

To enhance participants’ adherence, dedicated researchers will maintain contact with them via WeChat to provide reminders for antenatal visits and follow-up appointments. In addition, participants will be encouraged to report any concerning symptoms or events to researchers at any time.

#### Intervention Group: Induction at 39 Weeks

Participants in the intervention group will be scheduled for admission between 39 weeks 0 days and 39 weeks 4 days of gestation. Depending on gestational age at recruitment, IOL will be scheduled as close as feasible to the time of randomization, and no later than 7 days post randomization. Failed induction is defined as failure to enter active labor after 18 hours of amniotomy and intravenous oxytocin administration. In such cases, delivery will proceed by CS.

- Cervical ripening: For women with an unfavorable cervix (Bishop score <6, Supplementary Table 1), cervical ripening is required first. The preferred method is balloon catheter with early artificial rupture of membranes (ARM)(21), unless a specific contraindication or clinician/patient preference exists. Dinoprostone vaginal insertion is permitted as an alternative but are not the primary recommendation. Failure of cervical ripening is defined as a Bishop score remaining <6 after removal of the balloon catheter or dinoprostone. Subsequent management including repeat cervical ripening or caesarean section will be determined based on clinical assessment and patient preference.
- Direct labour induction: For women with a favorable cervix (Bishop score ≥6), Intravenous oxytocin infusion is the first-line method for IOL.

#### Control Group: Expectant Management

Participants in the control group will receive expectant management with weekly antenatal monitoring. Participants will be instructed to present to the hospital at any time if they experience regular uterine contractions (every 3-5 minutes, lasting over 30 minutes), rupture of membranes, vaginal bleeding exceeding normal menstrual volume, or decreased fetal movements.

Artificial intervention will not be initiated unless a medical indication emerges (e.g., suspected macrosomia, fetal distress, oligohydramnios, or gestational hypertension). Participants will be admitted upon the spontaneous onset of labour. If labour does not commence by 40 weeks 5 days, IOL will be performed using the same procedures as described for the intervention group.

#### Follow-up

Maternal and perinatal outcomes will be collected from delivery until hospital discharge. Infant follow-up visits are scheduled for 6 weeks and 6 months postpartum at the neonatal outpatient department to assess growth and neurodevelopment. Participants will remain in the study unless they withdraw consent or are discontinued by the Ethics Committee. For participants who deviate from the protocol but do not withdraw consent, outcome assessment will proceed as planned to enable ITT analysis.

#### Biological Sample Collection and Storing

For participants consenting to biological sampling, specimens will be collected at predefined intervals:

- Maternal: Vaginal secretions at 39 weeks’ gestation; maternal faeces;
- Infant: Cord blood and meconium at birth; neonatal faeces at day 2, week 1, and week 3; and infant faeces at 6 weeks, 3 months, and 6 months.

Cord blood samples will be processed into whole blood and plasma fractions and stored at −80°C. Stool samples collected in hospital will be immediately preserved on dry ice and transferred to −80°C storage. Samples collected at home will be transported to the hospital under cold-chain conditions within 2 hours and stored at −80°C upon arrival. These biological samples are stored for future exploratory analyses, including multi-omics analyses, and are not part of the primary outcomes of the present trial.

### Outcomes

The primary outcome is the caesarean rate, defined as the proportion of all randomized participants whose delivery is achieved via CS. The key secondary outcome is a composite of severe neonatal morbidity and perinatal mortality, including: antepartum/intrapartum/neonatal death; advanced respiratory support within first 72 hours of birth; Apgar score ≤ 3 at 5 minutes; neonatal encephalopathy; seizures; sepsis; pneumonia; meconium aspiration syndrome; birth trauma; intracranial hemorrhage or subgaleal hemorrhage; and hypotension requiring pressors (see detailed definition in Supplementary Table 2).

Other secondary outcomes (detailed in Supplementary Table 2) include:

- Maternal: Emergency/elective caesarean rates, operative vaginal delivery, gestational age at delivery, chorioamnionitis, grade III/IV perineal lacerations, postpartum haemorrhage/infection, ICU admission, and patient-reported pain (10-point Likert scale).
- Neonatal & Infant: Birth weight, macrosomia, large-for-gestational-age (LGA), small-for-gestational-age (SGA), neonatal acidosis, and shoulder dystocia. Early developmental outcomes will be assessed at 6 weeks and 6 months postpartum using WHO growth Z-scores, Bayley Scales of Infant Development (Bayley-4), and Ages and Stages Questionnaires (ASQ-3).
- Procedural: Epidural analgesia rate, maternal/neonatal length of stay, and hospitalisation costs.
- Exploratory Multi-omics: Longitudinal profiling of gut microbiota and metabolome (from meconium to 6 months), and cord blood proteomics/metabolomics. These biological signatures will be analysed and published independently to explore the systemic impact of IOL.

### Adverse Event

Adverse events (AEs) will be systematically captured via regular electronic case report form (eCRF) review, hospital electronic medical records (EMR), and non-systematically via participant reports. Serious adverse events (SAEs), including maternal/perinatal death or life-threatening complications, must be reported to the Ethics Committee and the Data and Safety Monitoring Committee (DSMC) within 24 hours. The hospital will provide immediate medical care for any study-related harms. Non-serious AEs will be recorded and summarized for the DSMC’s annual safety review.

### Sample size and recruitment capacity

#### Sample size calculation

The sample size was calculated using the Fleiss correction with normal approximation to binomial distribution(22). Based on the clinical data from our hospital, the overall CS rate for low-risk women is approximately over 30%. However, after excluding maternal request CS and assuming stricter standardized indications within this trial, we conservatively estimate the CS rate in expectant management group to be 16%. A risk difference of 6.5 percentage points (16% vs 9.5%) was considered clinically meaningful for the primary outcome, informed by preliminary observational data from our center. Accounting for a 5% protocol deviation rate (e.g., participants requesting advance or postpone the scheduled induction contrary to their allocation) and a two-sided alpha of 5% (α=0.05) with 80% power (1−β=0.80), a total of 1,020 participants (510 per group) is required. To account for a 5% loss to follow-up, the final sample size is set at 1,074 participants (537 per group). A sensitivity calculation of sample sizes under varying events and deviation rates is provided in Supplementary Table 3.

#### Recruitment capacity

With an annual delivery volume of approximately 28,000 and over 10 obstetricians participating, we anticipate a recruitment rate of 8-10 participants per week. We estimate that total recruitment will be completed within 28 months.

### Data collection, management and monitoring

#### Data collection and management

Clinical data will be systematically collected by certified obstetricians and paediatricians using the EMR and CRFs. Notably, all paediatricians involved are certified examiners for the Bayley-4 and ASQ-3 developmental scales. To ensure high data integrity, two research nurses will independently upload and verify all records within a web-based Electronic Data Capture (EDC) system, which maintains a comprehensive audit trail for all modifications.

While primary and perinatal outcomes are readily accessible via the EMR for all participants delivering at our facility, specific strategies have been implemented to minimize attrition for 6-month infant follow-up. These include providing maternal-infant counselling services up to 6 months postpartum and maintaining contact via a dedicated WeChat group. In cases of loss to follow-up for late secondary outcomes, baseline characteristics and earlier data will still be retrieved from the EMR to support the ITT analysis.

#### Data and Safety Monitoring Board (DSMB)

An independent DSMB, comprising experts in clinical medicine, biostatistics, and ethics who are unaffiliated with the sponsor or funder, will be established to oversee participant safety and trial integrity. The DSMB is responsible for conducting periodic reviews of safety data and providing recommendations regarding the continuation, modification, or termination of the trial. Prior to each meeting, the Biostatistics Coordinating Center will submit a report detailing participant demographics, protocol adherence (including deviations and mitigation actions), safety profiles (categorized by System Organ Class and Preferred Term codes), and site performance metrics such as recruitment progress and biospecimen collection rates.

#### Interim Analysis and Stopping Rules

A formal interim analysis is scheduled after 500 participants have complete data on the primary outcome. This analysis will evaluate the recruitment performance, participant retention, efficacy and safety profile of interventions, biospecimen collection rate. The DSMB may recommend early trial termination if the interim analysis demonstrates: (1) overwhelming evidence of efficacy (*p* < 0.002 with a favorable risk-benefit profile); (2) significant safety concerns, such as a disproportionately higher SAE rate in the intervention group; or (3) futility, where the primary endpoint is unlikely to be achieved. Any final decision regarding early termination must be proposed by the DSMB and formally approved by the Ethics Committee.

One blinded sample size re-estimation will be performed in this interim analysis. The re-estimation will be conducted by an independent statistician not otherwise involved in trial conduct. The reassessment will use the pooled observed event rate across treatment groups, without disclosure of arm-specific outcome rates or comparative treatment effect estimates to the study team. The clinically relevant treatment effect (risk difference of 6.5 percentage points), significance level, power, primary estimand, and final analysis method will remain unchanged. The total sample size will be recalculated using the same prespecified method at planning stage, replacing the design-stage pooled event-rate assumption with the interim pooled estimate. The total sample size may be increased up to a maximum of 1,200 but will not be reduced below the originally planned sample size.

## Statistical methods

Statistical analysis will follow the ITT principle, with per-protocol (PP) analysis as a supportive exploratory measure. All tests will be two-side (*p* < 0.05). A statistical analysis plan (SAP) will be finalized before the last enrollment. Baseline characteristics will be summarized using descriptive statistics to assess the balance between groups, with categorical data as counts (percentages) and continuous variables as mean ± SD or median (IQR) based on normality (Shapiro-Wilk normality tests and histograms).

The primary outcome (caesarean rate) will be analyzed using modified Poisson regression with robust variance estimation to estimate relative risk (RR) with 95% CIs, adjusting for stratification factors (age and parity). Adjusted risk differences (RDs) will be estimated using a linear probability model with robust standard errors, including stratification factors (age and parity) as covariates. Additional baseline characteristics may be controlled for in complementary analysis in case of baseline imbalances. Secondary outcomes will employ similar regressions for binary data, linear or quantile regression for continuous data, and Kaplan-Meier curves with log-rank tests for time-to-event data. To manage missing outcome data, multiple imputation and worst/best-case scenario simulations will be used. If attrition is substantial, inverse probability of censoring weighting (IPCW) will be used to mitigate attrition bias. If attrition is mainly attributable to pregnancy termination (e.g., stillbirth), competing risk analysis will be conducted. Exploratory subgroup analyses with interaction tests will be performed based on parity (nulliparous vs. multiparous), maternal age (<35 vs. ≥35 years), baseline bishop score (<6 vs. ≥6), BMI (<30 vs. ≥ 30 kg/m^2^). Finally, multi-modal integration of multi-omics data will be conducted and published as independent research.

### Ethics and dissemination

The study and its subsequent protocol amendment (approved 26 Jan 2026) have received ethical clearance from the Ethics Committee of Women’s Hospital, School of Medicine, Zhejiang University (initial approval: 10 Jun 2025). Any further modifications will be submitted for formal review. The trial was prospectively registered on ClinicalTrial.gov (NCT07082530). Participating obstetricians will obtain written informed consent from all eligible women or their legally authorized representatives prior to enrollment. A separate, optional consent will be required for biological sample donation for multi-omics analysis; refusal will not affect participation in the main trial.

Confidentiality is maintained via unique IDs; identifiers are stored separately with access restricted to the research team, Ethics Committee, and DSMB. De-identified data will be archived five years post-completion. Findings and multi-omics results will be published in peer-reviewed journals. De-identified data and codes are available from the corresponding author upon reasonable academic request.

### Patient and public involvement

The study design was informed by interviews with pregnant women over 39 weeks and a clinician survey, revealing anxiety over fetal overgrowth and induction risks. These insights confirmed the need for robust clinical evidence. Biospecimen collection protocols were optimized based on patient feedback; donation remains optional with guided self-collection. To ensure participant benefit, Bayley-4 and ASQ-3 reports will be provided to families after follow-up, ensuring a patient-centered approach to dissemination.

### Protocol version

Version 4.0, May 4, 2026. This revised protocol reflects key optimizations from earlier versions, including:

- Integration of multi-omics biospecimen sampling at multiple time points;
- Refined recruitment workflows to enhance participant engagement;
- Optimization of secondary outcomes to better evaluate offspring development and maternal psychological well-being.

## Discussion

Since the ARRIVE RCT in 2018(14), elective IOL at 39 weeks in low-risk pregnancies has gained increasing popularity in obstetric practice. However, consensus on its routine use has not yet been established in international guidelines. Further high-quality evidence across diverse populations is needed to confirm the generalizability of its findings. The LIRIC trial is designed to address this gap by evaluating the effects of 39-week induction on caesarean section rates and severe perinatal outcomes in a Chinese population.

Compared with previous studies, LIRIC incorporates several notable design features. First, both nulliparous and multiparous women are included, which may enhance the applicability of findings across broader obstetric populations. Second, follow-up of offspring is extended to 6 months postpartum, allowing assessment of early neurodevelopment using standardised tools, including the Bayley-4. In addition, follow-up results will be communicated to parents, accompanied by appropriate feeding and caregiving guidance, which may provide additional clinical value to participants. Third, the integration of longitudinal biological sampling with detailed clinical phenotyping enables exploratory analyses of potential biological pathways underlying observed outcomes. Furthermore, in light of evidence from SWEPIS and INDEX demonstrating increased risks associated with expectant management beyond 41 weeks(7, 8), contemporary clinical practice has shifted towards earlier intervention. Within this evolving landscape, it is timely to re-evaluate the potential benefits of induction at 39 weeks relative to the current expectant management window that ends before 41 complete weeks.

This study has several methodological strengths. The relatively large sample size is expected to ensure adequate statistical power for the primary outcome. Blinding of paediatric assessors and laboratory personnel is expected to reduce outcome assessment bias. Potential challenges include participant recruitment, adherence to the allocated interventions and loss to follow-up during infancy. Based on previous studies, a substantial proportion of eligible women may decline participation; however, recruitment will be supported by the high delivery volume across multiple hospital campuses. Public discourse on labour induction—particularly through social media—may influence participants’ preferences and affect adherence to the assigned strategy. To mitigate this, adherence will be promoted through patient education and regular follow-up. Additionally, analyses will primarily follow the ITT principle with PP analyses performed to assess the robustness of the findings and minimise the impact of protocol deviations. Loss to follow-up in postnatal assessments is also anticipated. Measures such as establishing dedicated communication channels (e.g., WeChat groups) will be implemented to facilitate participant engagement and ongoing support.

High caesarean section rates and adverse neonatal composite outcomes remain major concerns in obstetrics worldwide. In China, the caesarean section rate reached 48.57% in 2024(23), and substantial regional disparities in neonatal mortality persists (national average 2.5‰ vs 0.07‰ at the study center in 2024)(24). Against this backdrop, evaluating the benefits and potential risks of 39-week IOL across diverse settings is of considerable clinical importance. Given the complexity of conducting multicenter RCT, the LIRIC study will adopt a phased implementation strategy. The current protocol represents the first phase, focusing on evaluating the impact on caesarean section rates in a single-center setting and establishing a standardised biobank. In the second phase, the study will expand to a national multicenter design to further investigate the effects of 39-week IOL on severe maternal and neonatal outcomes. The findings of LIRIC will contribute to the existing evidence base on the optimal timing of delivery.

## Supporting information

SPIRIT 2025 Checklist

## Data Availability

All data produced in the present work are contained in the manuscript.

## Acknowledgements

The authors thank the obstetricians and patients for their valuable suggestions on the study design. We also acknowledge all participating physicians, research nurses and research assistants for their contributions to the study implementation, whose contributions will be fully recognised in subsequent outcome publications.

## Author Contributions

The guarantor is D.Z. and W.L., the co-primary investigators for LIRIC. All authors have contributed to the design of the study and have approved the final version of the protocol and manuscript.

## Competing interests

None declared.

## Funding

This study is supported by the National Key Research and Development Program of China (2025YFC2708200), the Ministry of Education Key Program for Basic and Interdisciplinary Sciences (JYB2025XDXM508), and the National Natural Science Foundation of China (82394424).

## Notes

### Competing Interest Statement

The authors have declared no competing interest.

### Clinical Trial

NCT07082530

### Author Declarations

The Ethics Committee of Women's Hospital, Zhejiang University School of Medicine gave ethical approval for this study (approval number: IRB-20250211-R).

## Reference

1. World Health Organization. WHO Global Survey on Maternal and Perinatal Health: Induction of labour data. Geneva; 2010.

2. Martin JA, Osterman MJK. Induction of Labor Increases in the United States: 2016 to 2024. NCHS Data Brief. 2026(554).

3. NHS English. NHS Maternity Statistics, England, 2024-25. London; 2025.

4. Australian Institute of Health and Welfare. Australia’s mothers and babies: Onset of labour. Canberra; 2026.

5. Zhu J, Xue L, Shen H, Zhang L, Lu D, Wang Y, et al. Labor induction in China: a nationwide survey. BMC Pregnancy Childbirth. 2022;22(1):463.

6. Middleton P, Shepherd E, Morris J, Crowther CA, Gomersall JC. Induction of labour at or beyond 37 weeks’ gestation. Cochrane Database Syst Rev. 2020;7(7):CD004945.

7. Keulen JK, Bruinsma A, Kortekaas JC, van Dillen J, Bossuyt PM, Oudijk MA, et al. Induction of labour at 41 weeks versus expectant management until 42 weeks (INDEX): multicentre, randomised non-inferiority trial. BMJ. 2019;364:344.

8. Wennerholm UB, Saltvedt S, Wessberg A, Alkmark M, Bergh C, Wendel SB, et al. Induction of labour at 41 weeks versus expectant management and induction of labour at 42 weeks (SWEdish Post-term Induction Study, SWEPIS): multicentre, open label, randomised, superiority trial. BMJ. 2019;367:6131.

9. World Health Organization. WHO recommendations on induction of labour at or beyond term. Geneva; 2022.

10. Obstetrics Subgroup CSoO, Gynecology CMA. Guideline of cervical ripening and labor induction during the third trimester pregnancy (2024). Zhonghua Fu Chan Ke Za Zhi. 2024;59(11):819–28.

11. National Institute for Health and Care Excellence. Inducing labour. NICE Guideline NG207. London; 2021.

12. Robinson D, Campbell K, Hobson SR, MacDonald WK, Sawchuck D, Wagner B. Guideline No. 432a: Cervical Ripening and Induction of Labour - General Information. J Obstet Gynaecol Can. 2023;45(1):35–44 e1.

13. Queensland Health. Induction of labour. Brisbane; 2022.

14. Grobman WA, Rice MM, Reddy UM, Tita ATN, Silver RM, Mallett G, et al. Labor Induction versus Expectant Management in Low-Risk Nulliparous Women. N Engl J Med. 2018;379(6):513–23.

15. Hong J, Atkinson J, Roddy Mitchell A, Tong S, Walker SP, Middleton A, et al. Comparison of Maternal Labor-Related Complications and Neonatal Outcomes Following Elective Induction of Labor at 39 Weeks of Gestation vs Expectant Management: A Systematic Review and Meta-analysis. JAMA Netw Open. 2023;6(5):e2313162.

16. Werner EF, Schlichting LE, Grobman WA, Viner-Brown S, Clark M, Vivier PM. Association of Term Labor Induction vs Expectant Management With Child Academic Outcomes. JAMA Netw Open. 2020;3(4):e202503.

17. Lindquist A, Hastie R, Kennedy A, Gurrin L, Middleton A, Quach J, et al. Developmental Outcomes for Children After Elective Birth at 39 Weeks’ Gestation. JAMA Pediatr. 2022;176(7):654–63.

18. Burger RJ, Mol BW, Ganzevoort W, Gordijn SJ, Pajkrt E, Van Der Post JAM, et al. Offspring school performance at age 12 after induction of labor vs non-intervention at term: A linked cohort study. Acta Obstet Gynecol Scand. 2023;102(4):486–95.

19. Chan AW, Boutron I, Hopewell S, Moher D, Schulz KF, Collins GS, et al. SPIRIT 2025 statement: updated guideline for protocols of randomised trials. BMJ. 2025;389:e081477.

20. Obstetrics Subgroup CSoOaG, Chinese Medical Association. Expert consensus on timing of termination of pregnancy for complications and comorbidities during pregnancy,. Chinese Journal of Obstetrics and Gynecology. 2020;55(10):649–58.

21. Jones MN, Palmer KR, Pathirana MM, Cecatti JG, Filho OBM, Marions L, et al. Balloon catheters versus vaginal prostaglandins for labour induction (CPI Collaborative): an individual participant data meta-analysis of randomised controlled trials. Lancet. 2022;400(10364):1681–92.

22. Fleiss JL. Statistical Methods for Rates and Proportions (2nd ed.): Wiley; 1981.

23. National Center for Healthcare Quality Management in Obstetrics. National Report on Maternal Health Care Service and Quality Safety. Beijing; 2026.

24. National Bureau of Statistics. 2024 Statistical monitoring report on the National Program for Child Development (2021–2030). Beijing; 2025.

