## Supplementary Material for "Labour Induction in low-risk women at 39 weeks of gestation: a Randomised trial in China (LIRIC) - Protocol of an open label, randomised controlled trial"

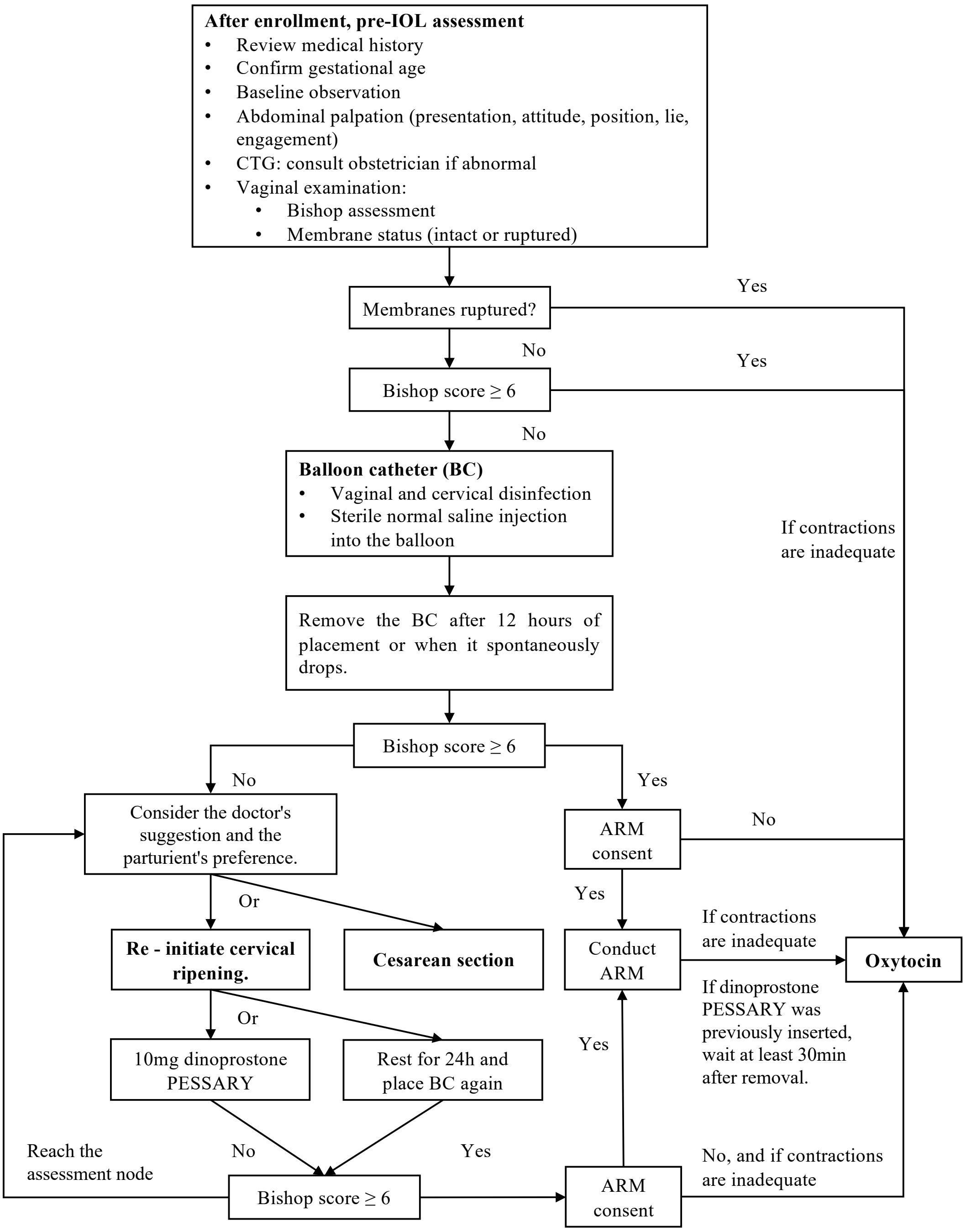


**Supplementary Fig. 1. Flow chart of induction of labor methods (post - randomization).**

Abbreviations: IOL, induction of labor; CTG, cardiotocography; BC, balloon catheter; ARM, artificial rupture of membrane.

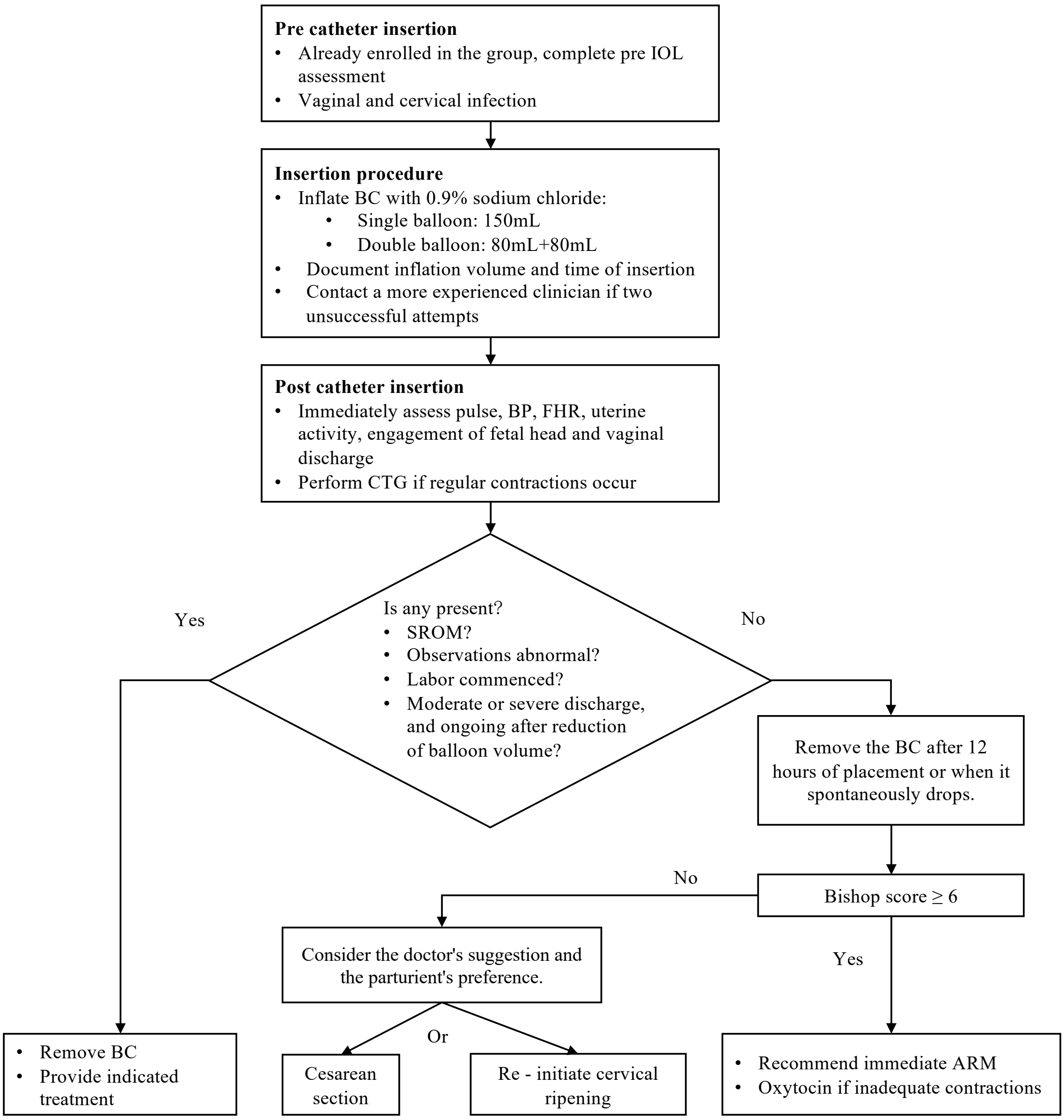


**Supplementary Fig. 2. Flow chart of balloon catheter.**

Abbreviations: IOL, induction of labor; CTG, cardiotocography; BC, balloon catheter; BP, blood pressure; FHR, fetal heart rate; SROM, spontaneous rupture of membranes; ARM, artificial rupture of membrane.

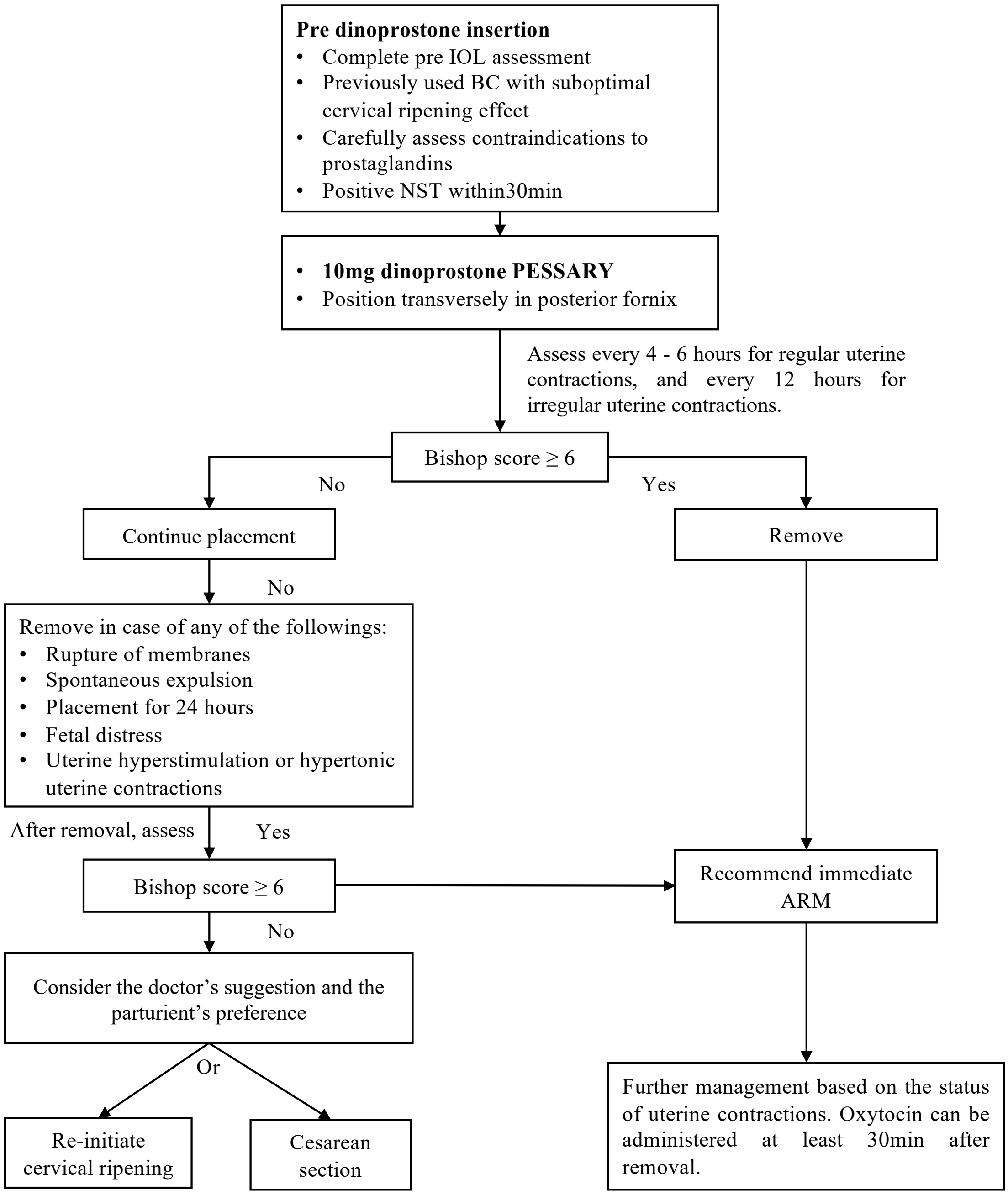


**Supplementary Fig. 3. Flow chart of dinoprostone insertion.**

Abbreviations: IOL, induction of labor; NST, Non-stress test; BC, balloon catheter; ARM, artificial rupture of membrane.

**Supplementary Table 1. Scoring System for the Bishop Score.**

| **Score** | 0 | 1 | 2 | 3 |
|---|---|---|---|---|
| Cervical Dilation (cm) | Unopened | 1-2 | 3-4 | >4 |
| Cervical Effacement (%) | 0-30 | 40-50 | 60-70 | ≥ 80 |
| Station of Presenting Part (in relation to ischial spines) | -3 | -2 | -1/0 | +1/+2 |
| Cervical Consistency | Hard | Medium | Soft | |
| Cervical Position | Posterior | Mid - position | Anterior | |

**Supplementary Table 2. Outcomes**

| **Primary outcome** | | |
|---|---|---|
| Outcome Measure | Measure Description | Time Frame |
| Incidence of cesarean section | The proportion of deliveries completed by cesarean section | Delivery day |
| **Key secondary outcomes** | | |
| Outcome Measure | Measure Description | Time Frame |
| Composite incidence of severe neonatal morbidity and perinatal mortality | The composite incidence of any of the following:<br>1. Antepartum, intrapartum, or neonatal death<br>2. Intubation, continuous positive airway pressure (CPAP) or high-flow nasal cannula (HFNC) for ventilation or cardiorespiratory support within first 72 hours<br>3. Apgar ≤ 3 at 5 minutes<br>4. Neonatal encephalopathy<br>5. Seizures<br>6. Sepsis (presence of a clinically ill infant in whom systemic infection is suspected with a positive blood, cerebral spinal fluid (CSF), or catheterized/suprapubic urine culture; or, in the absence of positive cultures, clinical evidence of cardiovascular collapse or an unequivocal X-ray confirming infection).<br>7. Pneumonia confirmed by X-ray or positive blood culture.<br>8. Meconium aspiration syndrome<br>9. Birth trauma (bone fractures, brachial plexus palsy, other neurologic injury, retinal hemorrhage, facial nerve injury)<br>10. Intracranial hemorrhage or subgaleal hemorrhage<br>11. Hypotension requiring pressor support | Hospital discharge |
| **Maternal secondary outcomes** | | |
| Outcome Measure | Measure Description | Time Frame |
| Emergency Cesarean Rate | Cesarean delivery performed immediately or shortly after clinical decision due to imminent threat to the life of the mother or fetus | Delivery day |
| Category 1 Emergency Cesarean Rate | Cesarean performed within approximately 30 minutes due to immediate life-threatening conditions (e.g., uterine rupture, severe fetal distress) | Delivery day |
| Category 2 Emergency Cesarean Rate | Cesarean performed within 60–75 minutes due to urgent but not immediately life-threatening indications (e.g., progressive fetal distress, labor arrest) | Delivery day |
| Elective Cesarean Rate | Planned cesarean performed at or after 39 weeks in | Delivery day |

| | | |
|---|---|---|
| | the absence of emergency conditions, meeting all of the following:<br>1. Surgery scheduled ≥24 hours before onset of labor;<br>2. No spontaneous contractions or rupture of membranes;<br>3. Clear medical indications (e.g., placenta previa, breech presentation) or maternal request. | |
| Operative Vaginal Delivery | The rate of vaginal deliveries assisted with instruments such as forceps or vacuum. | Delivery day |
| Gestational Age at Delivery | Expressed in weeks and decimal days (e.g., 39 weeks + 5 days = 39.7 weeks). | Delivery day |
| Chorioamnionitis | Incidence of chorioamnionitis, defined as a clinical diagnosis before delivery as: maternal body temperature rises (>38°C) with 2 or more of the following symptoms:<br>1. Maternal tachycardia (>100 beats/min);<br>2. Fetal tachycardia (>160 beats/min)<br>3. Uterine tenderness and irritability<br>4. Purulent or malodorous amniotic fluid or vaginal discharge<br>5. Maternal leukocytosis (white blood cell count > 15000/mm3) | Delivery day |
| Third-or fourth-degree perineal laceration | The incidence of third-or fourth-degree perineal laceration | Delivery day |
| Postpartum hemorrhage | Refer to the ARRIVE cohort, defined as any of the following situations:<br>1. Transfusion<br>2. Non-elective hysterectomy<br>3. Use of two or more uterotonics other than oxytocin<br>4. Other surgical interventions such as uterine compression sutures, uterine artery ligation, embolization and hypogastric ligation, balloon tamponade<br>5. Curettage | Hospital discharge |
| Postpartum infection | Refer to the ARRIVE cohort, defined as any of the following situations:<br>1. Clinical diagnosis of endometritis<br>2. Wound reopened for hematoma, seroma, infection or other reasons<br>3. Cellulitis requiring antibiotics<br>4. Pneumonia<br>5. Pyelonephritis<br>6. Bacteremia unknown source<br>7. Septic pelvic thrombosis | Hospital discharge |

| | | |
|---|---|---|
| ICU admission | The proportion of admission to the ICU | Hospital discharge |
| Maternal death | The incidence of maternal death | Hospital discharge |
| Preeclampsia/gestational hypertension | The incidence of preeclampsia or gestational hypertension diagnosed during labor | Hospital discharge |
| Maternal venous thromboembolism | Deep vein thrombosis or pulmonary embolism | Hospital discharge |
| Maternal pain | Median patient-reported pain outcomes with a 10-point Likert scale | Hospital discharge |
| **Fetal and neonatal secondary outcomes** | | |
| Outcome Measure | Measure Description | Time Frame |
| Birth weight | The mean birth weight | Delivery day |
| Female gender | The proportion of female offspring in terms of physiological gender | Delivery day |
| Macrosomia | Birth weight > 4000g | Delivery day |
| Incidence of large for gestational age | According to the infant's gender and gestational age, the birth weight exceeds the 90th percentile of infants of the same gestational age | Delivery day |
| Incidence of small for gestational age | According to the infant's gender and gestational age, the birth weight is less than the 10th percentile of infants of the same gestational age | Delivery day |
| Apgar scores at 5 min | Apgar score at 5 minutes after birth | Delivery day |
| Neonatal acidosis | The incidence of neonatal acidosis defined as umbilical artery blood pH < 7.10 | Delivery day |
| Shoulder dystocia | The incidence of shoulder dystocia | Delivery day |
| Hyperbilirubinemia | The incidence of hyperbilirubinemia requiring phototherapy or exchange transfusion | Hospital discharge |
| Transfusion of blood products or blood | The incidence of neonatal transfusion of blood products or blood | Hospital discharge |
| Hypoglycemia incidence | The incidence of hypoglycemia (blood glucose < 35mg/L) requiring intravenous treatment | Hospital discharge |
| Neonatal intensive care unit (NICU) admission | The incidence rate of NICU admission lasting more than 48 hours | Hospital discharge |
| **Infant early-development outcomes** | | |
| Outcome Measure | Measure Description | Time Frame |
| Physical measurement indicators | Infant weight, length, head circumference, chest circumference, sitting height, upper arm circumference, subcutaneous fat, height-chest circumference index, crown-rump length/length | 6 weeks and 6 months |
| Growth balance indicators | Weight-for-length, length-for-age, weight-for-age z scores of infants, calculated using the WHO tool WHO Anthro Survey Analyser | 6 weeks and 6 months |

| | | |
|---|---|---|
| Growth retardation | Height standard deviation for age < mean-2S or percentile method < P3 | 6 weeks and 6 months |
| Abnormal neurological examination | The proportion of abnormal results in neurological examinations (hearing development, nerve reflexes) | 6 weeks and 6 months |
| Bayley-4 scores of the Infant Development Scale | Total score, cognitive scale score, language scale score, motor scale score, social-emotional scale score, adaptive behavior scale score | 6 weeks and 6 months |
| ASQ-3 scores of the Ages and Stages Questionnaires | Total score, communication domain score, gross motor domain score, fine motor domain score, problem-solving domain score, personal-social domain score | 6 weeks and 6 months |
| Incidence of allergic diseases | Allergic diseases diagnosed by neonatologists, including cow's milk allergy, allergic rhinitis, eczema/allergic dermatitis, asthma, wheezing and other allergic disorders | 6 weeks and 6 months |
| **Multi-omics outcomes** | | |
| Outcome Measure | Measure Description | Time Frame |
| Gut microbiota development | Microbiota richness, dominant species, species transferred from mother to child, sub-group functional trajectory changes of the dominant species in offspring, and the transmissibility of species number and function | Meconium, first 48h, 7 days, 3 weeks, 6 weeks, 3 months and 6 months |
| Fecal metabolomic characteristics | Non-targeted metabolomic macroscopic differences, core metabolomic differences (including tryptophan, fatty acids), and differences in gut microbiota metabolites | Meconium, first 48h, 7 days, 3 weeks, 6 weeks, 3 months and 6 months |
| Cord blood metabolomic characteristics | Assessment of non-targeted metabolomic macroscopic differences and core metabolomic differences (including tryptophan, fatty acids) in cord blood plasma | Delivery day |
| Cord blood proteomic characteristics | Analysis of the proteomic characteristics of cord blood whole blood to trace back the health status during the fetal period | Delivery day |
| **Utilization of Medical Resources** | | |
| Outcome Measure | Measure Description | Time Frame |
| Use of epidural analgesia | Rate of epidural analgesia use | Delivery day |
| Maternal length of hospital stays | Median maternal hospital stays, measured in hours | Delivery day |
| Neonatal length of hospital | Median neonatal hospital stays, measured in hours | Delivery day |

| | | |
|---|---|---|
| stays | | |
| Pre-insurance hospitalization cost | Total hospitalization cost during delivery, before insurance reimbursement | Delivery day |
| Out-of-pocket hospitalization cost | Out-of-pocket cost incurred during delivery hospitalization | Delivery day |

**Supplementary Table 3. Sample Sizes per Group for Different Primary Outcome Rates**

| Nominal Primary Outcome Rate in Induction Group | Nominal Primary Outcome Rate in Expectant Management Group | | | | | |
|---|---|---|---|---|---|---|
| | 13% | 14% | 15% | 16% | 17% | 18% |
| 0% not delivering per protocol | | | | | | |
| 8% | 589 | 426 | 325 | 258 | 211 | 177 |
| 9.50% | 1279 | 803 | 557 | 413 | 320 | 257 |
| 11% | 4144 | 1907 | 1109 | 733 | 524 | 396 |
| 2.5% not delivering per protocol | | | | | | |
| 8% | 653 | 472 | 361 | 286 | 234 | 196 |
| 9.50% | 1417 | 890 | 617 | 457 | 355 | 285 |
| 11% | 4591 | 2113 | 1229 | 812 | 581 | 439 |
| 5% not delivering per protocol | | | | | | |
| 8% | 728 | 526 | 402 | 319 | 261 | 219 |
| 9.50% | 1579 | 992 | 688 | 510 | 395 | 317 |
| 11% | 5116 | 2355 | 1369 | 905 | 647 | 490 |
| 7.5% not delivering per protocol | | | | | | |
| 8% | 816 | 590 | 451 | 358 | 293 | 245 |
| 9.50% | 1770 | 1112 | 771 | 571 | 443 | 356 |
| 11% | 5735 | 2640 | 1535 | 1014 | 726 | 549 |
